# Maternal obesity shapes the B lymphocyte and antibody repertoires of human colostrum

**DOI:** 10.1101/2023.09.01.23294956

**Authors:** Erick Sánchez-Salguero, Diana Bonilla-Ruelas, Mario René Alcorta-García, Víctor Javier Lara-Diaz, Claudia Nohemí López-Villaseñor, Marion E G Brunck

## Abstract

The prevalence of obesity is rapidly increasing worldwide and its impact on future generations must be assessed. We recently showed that colostrum from mothers with obesity contained a significantly reduced B lymphocytes (CD19^+^) fraction. Here, in a subsequent transversal cohort study of 48 mothers, we exhaustively characterize the B lymphocytes subsets present in peripheral blood and colostrum from obese mothers and describe a pervasive alteration of the B lymphocytes compartment of human colostrum accompanied by a dysregulated antibody composition. We describe significant decreases in regulatory B cells and soluble IgA concentrations, combined with increases in soluble IgG and double negative 2 (CD19^+^, CD27^-^, IgD^-^, CD38^-^, CD24^-^, CD21^-^, CD11c^+^) B lymphocytes. These alterations correlated with maternal BMI and corporal fat %. We provide evidence for possibly autoimmune IgG present in obese colostrum, and for the proinflammatory consequences of obese colostrum *in vitro*. Beyond the impact of obesity, we evidence the selective presence of B lymphocyte subtypes in colostrum and *in situ* production of IgG antibodies, which expands our current understanding of the origin of colostrum IgG. As maternal milk antibodies play a crucial role in regulating neonatal gut immune development, this work uncovers maternal obesity as a potential risk factor for compromised breastmilk immune components, calling for more research on the long-term health of lactating infants.

## 1. INTRODUCTION

Tolerizing responses in the gut allow the establishment of the microbiota and efficient food digestion, contributing to health. Disruptions in these tolerizing responses cause inflammation that promote disease. For example, in ulcerative colitis, immune tolerance to commensal microbes is impaired^1^. This prevents the expansion of a tolerizing RORγ^+^ Treg population, fueling gut inflammation that primes the disease^2^. Interestingly, RORγ^+^Treg populations essential to the establishment of the microbiota are transmitted exclusively through breastmilk and persist through adulthood^3^. Therefore, essential immune responses in the adult gut are imprinted at least in part during breastfeeding and inappropriate responses prime adverse conditions later in life^4^.

Maternal obesity is a rising condition worldwide that correlates with variations in multiples breastmilk components including the microbiota, Human-milk oligosaccharides (HMO) and lipids^5–8^. However, reports on immune bioactives like cytokines and leukocytes remain scarce^9–12^. We recently evidenced a significant reduction of the B lymphocytes compartment in the colostrum of mothers with obesity^13^. Here, we further characterize 18 B lymphocytes subpopulations in obese colostrum and describe pervasive alterations of the resolved populations, including less B_reg_-like and more of a recently described pro-inflammatory B lymphocyte population, also known as double-negative 2 (DN2) B cells^14,15^. These alterations at the cellular level are accompanied by significant regulations in colostrum antibodies, including less soluble IgA (sIgA), and more soluble IgG (sIgG). Interestingly, obese colostrum sIgG had increased recognition of N-acetylglucosamine (GlcNAc) which is present on bacterial and fungal cell walls, but also composes various human tissues, hinting toward a possible transfer of autoimmunity^16–18^. Finally, we show that in contrast to colostrum from “lean” mothers, obese colostrum leads to activation of human macrophages *in vitro*. Overall, we describe here that maternal obesity regulates B lymphocytes subsets and antibodies in human colostrum, with possible long-lasting impact on the suckling neonate’s health.

## 2. MATERIALS AND METHODS

### 2.1 Human samples

This cross-sectional study was approved by the Ethics Committee of the Hospital Regional Materno Infantil, Servicios de Salud de Nuevo León, and by the IRB at the School of Medicine and Health Sciences, TecSalud, in Monterrey, Mexico (CarlMicrobio2018, Reg. No. DEISC-19 01 18 09). Eligible women attending the hospital for delivery were recruited between September 2022 and April 2023. Participation in the study was based on the following inclusion criteria: (1) maternal age between 18 and 34 years, (2) over 5 prenatal visits without any adverse event during pregnancy, (3) pre-pregnancy BMI >18.5 and <25, or >30, (4) term infant, and (5) willingness to participate. Exclusion criteria included (1) having received antibiotics anytime during the 3-month period before birth, or having received a prolonged antibiotic treatment (>3 months) anytime during pregnancy, (2) having received immunosuppressive doses of steroids during pregnancy, (3) history of any monoclonal antibody treatment, (4) history of chronic disease (outside of obesity), (5) suffering from any nutrition-related disease or dietary restrictions, (6) episodes of diarrhea during the last 2 weeks of pregnancy, (7) history of surgery within 12 months prior to pregnancy, (8) history of antineoplastic treatment. Elimination criteria included (1) having received antibiotics for >24 h post-birth, (2) newborn admission to NICU, (3) any additional cause impeding sample collection. Oxytocin was not used during labor. Pre-pregnancy weight was recalled, current height was measured and current body fat percentage (BF%) obtained by impedance at the time of recruitment. Additional variables collected or measured in this work included maternal age, primiparity, infant gender, gestational age at birth, mode of delivery, weight of infant at birth, volume of colostrum obtained, frequency of B lymphocytes subpopulations in blood and colostrum samples, type and concentrations of antibodies and frequency of antibody-producing cells in colostrum. Additional details are included in the study’s STROBE statement (Supplementary Table 1). Participants were allocated to the “obese” (BMI>30, BF%>30) or “lean” (18.5>BMI<25, BF%<22) groups according to the WHO guidelines. Upon recruitment and signed informed consent, 4 ml of maternal peripheral blood were drawn into K_2_EDTA coated tubes (BD Vacutainer®, cat. 366643). The same day, after infant feeding, the nipple area of the breast was gently cleaned with neutral soap and water, and 1-3 ml of colostrum were collected using a manual pump. Samples were immediately stored on ice until processing and all samples were processed within 2 h of collection.

### 2.2 Isolation of PBMC from peripheral blood

Ficoll-Paque (Fisher scientific, cat. 17-1440-03) was used to enrich PBMC from peripheral blood and residual erythrocytes were lysed using Pharm Lyse Solution (BD, cat. 555899), as per manufactureŕs instructions. Cells were manually counted using a Neubauer chamber, using 0.4% Trypan blue to discriminate dead cells). Three million live PBMC were then stained for flow cytometry in a total volume of 100 µl.

### 2.3 Cell enrichment from colostrum

Approximately 2 ml of colostrum were processed for cell enrichment prior to staining for flow cytometry, as previously reported^13,19^. Briefly, colostrum volumes were recorded, and samples were centrifuged at 400 rcf for 15 min at 4°C. Supernatant were stored away, and cell pellet were washed twice with PBS/1% FBS/2mM EDTA. Cells were manually counted using a Neubauer chamber using 0.4% Trypan blue to discriminate dead cells. Two million live cells were then stained for flow cytometry in a total volume of 100 µl.

### 2.4 Flow cytometry

The same conjugated monoclonal antibodies were used to stain both tissue types, and antibody and viability stains were titrated independently for each tissue. PBMC were staining with 1.38 µl of CD19-PerCP Cy5.5 (BioLegend, catalog number 302230), 5 µl of CD21-PE/DazzleTM 594 (BioLegend, catalog number 354922), 5 µl of CD24-Brilliant Violet 786TM (BioLegend, catalog number 311142), 2 µl of CD27-APC (BioLegend, catalog number 302810), 2.5 µl of CD38-Alexa Fluor® 700 (BioLegend, catalog number 397206), 2 µl of IgA-VioGreen (Miltenyi Biotec, catalog number 130-113-481), 3 µl of IgG APC Cy7 (BioLegend, catalog number 410732), 3 µl of IgD-VioBlue (Miltenyi Biotec, catalog number 130-123-258), and 5 µl of CD11c-Brilliant Violet 605TM (BioLegend, catalog number 744436), and 1 µl of Zombie Green (BioLegend, catalog number 423111); in a final volume of 100 µl of PBS/1% FBS/2mM EDTA. As previous publications suggested we optimized staining times for optimal resolution, and incubated 90 min at 4°C in the dark^20^.

For colostrum cells, 2x10^6^ cells were stained using a cocktail including 1.38 µl CD19-PerCP Cy5.5, 5 µl CD21-PE/DazzleTM 594, 5 µl CD24-Brilliant Violet 786TM, 2 µl CD27-APC, 3 µl CD38-Alexa Fluor® 700, 5 µl IgA-VioGreen, 5 µl IgG- APC Cy7, 5 µl IgD-VioBlue, 5 µl IgG-APC Cy7, and 5 µl CD11c-Brilliant Violet 605TM; in a final volume of 100 µl of PBS/1% FBS/2mM EDTA. After incubation, samples were washed and resuspended in PBS/1% FBS/2mM EDTA for immediate acquisition on a BD® FACSCelesta flow cytometer fitted with 405 nm, 488 nm, and 633 nm lasers and operated through the BD® FACSDiva software v.8. Compensation controls were used at each acquisition using compensation beads following manufacturer’s recommendations, and automatic compensation was performed prior to acquisition. Over 10^6^ events were recorded from each sample, with the FSC threshold adjusted to 50,000 or 5,000 for blood and colostrum, respectively. Analysis was performed using FlowJo X 10.0.7r2. The gating strategy initially optimized was based on previous reports and CD45 stained samples but FMO controls were used to adjust gates for both sample types^21,22^. The gating strategy is described in Supplementary Fig. 1.

### 2.5 IgM, sIgA and sIgG ELISA

Flat-bottom 96-well polystyrene plates were coated with 1:5000 PBS-diluted mouse anti-human monoclonal antibody for either IgM (Abcam, cat. ab200541), IgA (Abcam, cat. ab7400b) or IgG (Abcam, cat. ab72528), and incubated 12 h at 4 °C.

After blocking, dilutions of plasma or colostrum supernatants were incubated for 2 h at 37 °C. For detection, anti-human IgG, IgA, and IgM coupled to HRP (Abcam, cat. ab102420) were added at 1:8000 dilution and incubated for 1 h at 37 °C. Fifty µl/ well of TMB (Abcam, cat. ab171523) were then incubated 2 min. The reaction was stopped with 50 µl of 0.2 M H_2_SO_4_, and plates were read at 450 nm on a spectrophotometer (Tecan’s Magellan® universal reader). Quantitative standard curves were obtained for each isotype using serial dilutions from recombinant human IgA (Abcam, cat. ab91025), IgG (Abcam, cat. Ab91102) or IgM (Abcam, cat. Ab91117).

### 2.6 IgM, IgG, and IgA-secreting cells ELISPOT

Briefly, 96-well plates were covered with PVDF membranes. After methanol activation, membranes were coated with a 1:2500 dilution of mouse anti-human monoclonal antibody recognizing IgM, IgA, or IgG (Abcam, cats. ab200541, ab7400, ab72528, respectively). The plates were incubated for 12 h at 4° C, then blocked for 90 min at 25 °C. Colostrum-enriched cells or blood PBMC were seeded at 200,000 cells/well in RPMI 1640 with 10% FBS and 100 U/ml penicillin, 0.1 mg/ml streptomycin. Plates were incubated for 18 h at 37 °C and 5% of CO_2_. For detection, a 1:10,000 dilution of HRP-conjugated goat anti-human IgG, IgA, and IgM (Abcam, cat. ab102420) was incubated 1 h at 37°C. Finally, 50 µl/well 3,3′- DAB (Sigma-Aldrich, cat. D4418) were added. Membranes were then washed and dried, and pictures were acquired with a Stereoscopic Microscope (Nikon, cat. SMZ1500). Spots were counted using the Analyze Particles command in ImageJ (Java®).

### 2.7 Colostrum-mediated macrophage cytokine production

To produce human macrophage-like cells, U937 cells (ATCC, CRL-1593.2) were differentiated over 24 h using 10 ng/ml PMA (Sigma-Aldrich®, cat. 79346) in RPMI 1640, supplemented as above. After 24 h, the media was replaced without PMA but including 2.5% 0.22 µm-filtered colostrum supernatant. After a further 24 h, cells were washed, fresh media without colostrum was added. To quantify cytokines, 5 µL of culture supernatants were obtained every 2 h for a total of 16 h. Samples were centrifuged and stored at -20 °C until quantification. Cytokines were quantified using a LEGENDplex kit (BioLegend, cat. 740808) according to the manufacturer’s instructions. All samples were immediately read on a BD® FACSCelesta flow cytometer fitted with 405 nm, 488 nm, and 633 nm lasers and operated through the BD® FACSDiva software version 8. Analysis and quantitative data were obtain using the FCAP Array software v 3.0 SoftFlow® LEGENDplex™ Cloud-based Data Analysis Software online^23^.

### 2.8 Statistical Analysis

Shapiro-Wilk normality tests were performed on each B lymphocyte subtype dataset. Non-normal datasets were compared using Mann-Whitney or one-way Kruskal-Wallis test when comparing >2 groups. Means of normally distributed data were compared using Student’s t test. Correlations were investigated using Pearson rank or Spearman r. All these tests were performed in GraphPad Prism v.8 (GraphPad Software Inc®, San Diego CA, USA). Median (IQR) were compared using Independent-Samples Median test in SPSS v.26.

## 3. RESULTS AND DISCUSSION

### 3.1 B lymphocytes subpopulations are selectively present in colostrum

We recruited a total of 48 mothers to participate in this study. As per study design, the BMI and BF% of the cohort of mothers with obesity were significantly larger compared to the cohort of “lean” mothers, while no difference were observed in possible confounders such as maternal age, infant gender, gestational age, or delivery type between the groups (Table 1).

**Table 1:** Participants characteristics.

|  | Group |  |  |  |  |
| --- | --- | --- | --- | --- | --- |
| Variable | Lean, n = 23 |  | Obese, n = 25 |  | P value |
| Maternal age (years) <sup>a</sup> | 21 | (20 - 24) | 23.5 | (20 -27.5) | 0.099 |
| BMI (kg/m<sup>2</sup>) <sup>a</sup> | 21.4 | (18.7 - 24.1) | 30.3 | (30.0 - 31.9) | **<0.0001** |
| Body fat percentage (%) <sup>a</sup> | 20 | (15 - 22) | 39.1 | (34 - 42) | **<0.0001** |
| Gestational age (weeks) <sup>a</sup> | 38 | (37 - 39) | 38 | (37 - 39) | 0.690 |
| Primiparity <sup>a</sup> | 2 | (1 - 3) | 3 | (1.75 - 4) | 0.104 |
| Vaginal delivery <sup>b</sup> | 23 | (100) | 24 | (92.3) | 0.491 |
| Newborn female gender <sup>b</sup> | 9 | (39.1) | 15 | (60) | 0.156 |
| Newborn male gender <sup>b</sup> | 14 | (60.9) | 10 | (40) |  |
| Birth weight (g) <sup>a</sup> | 3070 | (2700 - 3290) | 3000 | (2742 - 3255) | 0.411 |
| Birth height (cm) <sup>a</sup> | 49 | (47 - 50) | 49 | (47 - 50) | 0.832 |

Notes: BMI = Body Mass Index; a = Values expressed as median (IQR), statistical analysis with Independent-Samples Median test, b = Values expresses as frequency (%), statistical analysis with Fisher's exact test.

We applied an optimized 10-colour flow cytometry panel to detect 18 subpopulations of B lymphocytes in peripheral blood and colostrum. The gating strategy was based on classical and more recent markers used to subtype peripheral blood B lymphocytes (Table 2). We found a reduced fraction of the total B lymphocytes population in obese colostrum compared to the lean cohort (Supplementary Fig. 2), consistent with previous findings^13^. The reduction was not resumed in peripheral blood, suggesting that obesity regulates this compartment locally, with observed changes in colostrum.

**Table 2:** Flow cytometry-based identification of B lymphocyte subtypes.

| B cell subpopulation<br>(acronym) | Surface markers phenotype | References |
| --- | --- | --- |
| Total B cells (**B cells**) | CD19<sup>+</sup> | 27 |
| Transitional B cells type 1 (**T1**) | CD19<sup>+</sup>, CD27<sup>-</sup>, CD38<sup>hi</sup>,<br>CD24<sup>hi</sup>, CD21<sup>neg/low</sup> | 68,69 |
| Transitional B cells type 2 (**T2**) | CD19<sup>+</sup>, CD27<sup>-</sup>, CD38<sup>hi</sup>,<br>CD24<sup>hi</sup>, CD21<sup>+</sup> | 69,70 |
| Resting naïve B cells (**resN**) | CD19<sup>+</sup>, CD27<sup>-</sup>, IgD<sup>+</sup>, CD38<sup>+</sup>,<br>CD24<sup>-</sup>, CD11c<sup>-</sup> | 71,72 |
| Active naïve B cells (**actN**) | CD19<sup>+</sup>, CD27<sup>-</sup>, IgD<sup>+</sup>, CD38<sup>+</sup>, CD24<sup>-</sup>, CD11c<sup>+</sup> | 72,73 |
| Total plasma-like cells (**Plasma-like cells**) | CD19<sup>+</sup>, CD27<sup>hi</sup>, CD38<sup>hi</sup> | 74,75 |
| Plasma-like cells expressing IgA (**IgA<sup>+</sup> plasma-like cells**) | CD19<sup>+</sup>, CD27<sup>hi</sup>, CD38<sup>hi</sup>, IgA<sup>+</sup> |  |
| Plasma-like cells expressing IgG (**IgG<sup>+</sup> plasma-like cells**) | CD19<sup>+</sup>, CD27<sup>hi</sup>, CD38<sup>hi</sup>, IgG<sup>+</sup> |  |
| Unswitched memory B cells (**USwM**) | CD19<sup>+</sup>, CD27<sup>+</sup>, IgD<sup>+</sup> | 76,77 |
| IgG switched memory (**IgG<sup>+</sup> SwM**) | CD19<sup>+</sup>, CD27<sup>+</sup>, IgD<sup>-</sup>, IgG<sup>+</sup> |  |
| IgA switched memory (**IgA<sup>+</sup> SwM**) | CD19<sup>+</sup>, CD27<sup>+</sup>, IgD<sup>-</sup>, IgA<sup>+</sup> |  |
| Unswitched non-classical memory (**M CD27<sup>-</sup> IgD<sup>+</sup>**) | CD19<sup>+</sup>, CD38<sup>neg/low</sup>, CD24<sup>+</sup>, CD27<sup>-</sup>, IgD<sup>+</sup> | 78,79 |
| Switched non-classical memory or (**M CD27<sup>-</sup> IgD<sup>-</sup>**) | CD19<sup>+</sup>, CD38<sup>neg/low</sup>, CD24<sup>+</sup>, CD27<sup>-</sup>, IgD<sup>-</sup> |  |
| Double negative B cell subtype 1 (**DN1-like**) | CD19<sup>+</sup>, CD27<sup>-</sup>, IgD<sup>-</sup>, CD38<sup>-/+</sup>, CD24<sup>-</sup>, CD21<sup>+</sup>, CD11c<sup>-</sup> | 40,80 |
| Double negative B cell subtype 2 (**DN2-like**) | CD19<sup>+</sup>, CD27<sup>-</sup>, IgD<sup>-</sup>, CD38<sup>-</sup>, CD24<sup>-</sup>, CD21<sup>-</sup>, CD11c<sup>+</sup> |  |
| Double negative B cell subtype 3 (**DN3-like**) | CD19<sup>+</sup>, CD27<sup>-</sup>, IgD<sup>-</sup>, CD38<sup>-/+</sup>, CD24<sup>-</sup>, CD21<sup>-</sup>, CD11c<sup>-</sup> |  |
| Double negative B cell subtype 4 (**DN4-like**) | CD19<sup>+</sup>, CD27<sup>-</sup>, IgD<sup>-</sup>, CD38<sup>-</sup>, CD24<sup>-</sup>, CD21<sup>+</sup>, CD11c<sup>+</sup> |  |
| B reg-like cells (**B<sub>reg</sub>-like cells**) | CD19<sup>+</sup>, CD38<sup>+</sup>, CD24<sup>+</sup> | 81,82 |

In peripheral blood, we could detect all targeted subpopulations including rare subtypes like B_reg_-like and transitional B cells (Supplementary Fig. 3) with proportions consistent with previous reports^22,24–26^. In contrast, in colostrum multiple subtypes could not be detected, notably the early stages of B cell ontogeny including transitional and naïve B cells (Fig. 1A). In comparing B lymphocyte subtypes between blood and colostrum, we considered relative proportions (Supplementary Fig. 3), but also concentrations in original samples (Fig. 1) calculated from measured sample volumes, manual cell counts and % populations, to account for discrepancies in cellularity between blood and colostrum (Fig. 1). Differences were consistent between population % and concentrations. While early ontogeny B cells were absent from colostrum, differentiated subtypes were very significantly increased in this tissue, including B_reg_-like, DN2-like and plasma-like cells that were rare in peripheral blood (Fig. 1B), although definitive labelling should be based on functional assays such as cytokine and antibody production. While switched memory (SwM) B cells were present in similar concentrations in both colostrum and blood, unswitched memory B cells (USwM) were significantly underrepresented in colostrum compared to peripheral blood (Fig. 1C). Overall, the results describe pervasive, significant differences in % and concentrations of B lymphocytes subtypes between blood and colostrum. This may suggest a selective migration to the mammary acini and colostrum. It further describes human colostrum as containing multiple subpopulations of differentiated B cells, enriching the current state-of-the art^27,28^.

**Fig. 1:**
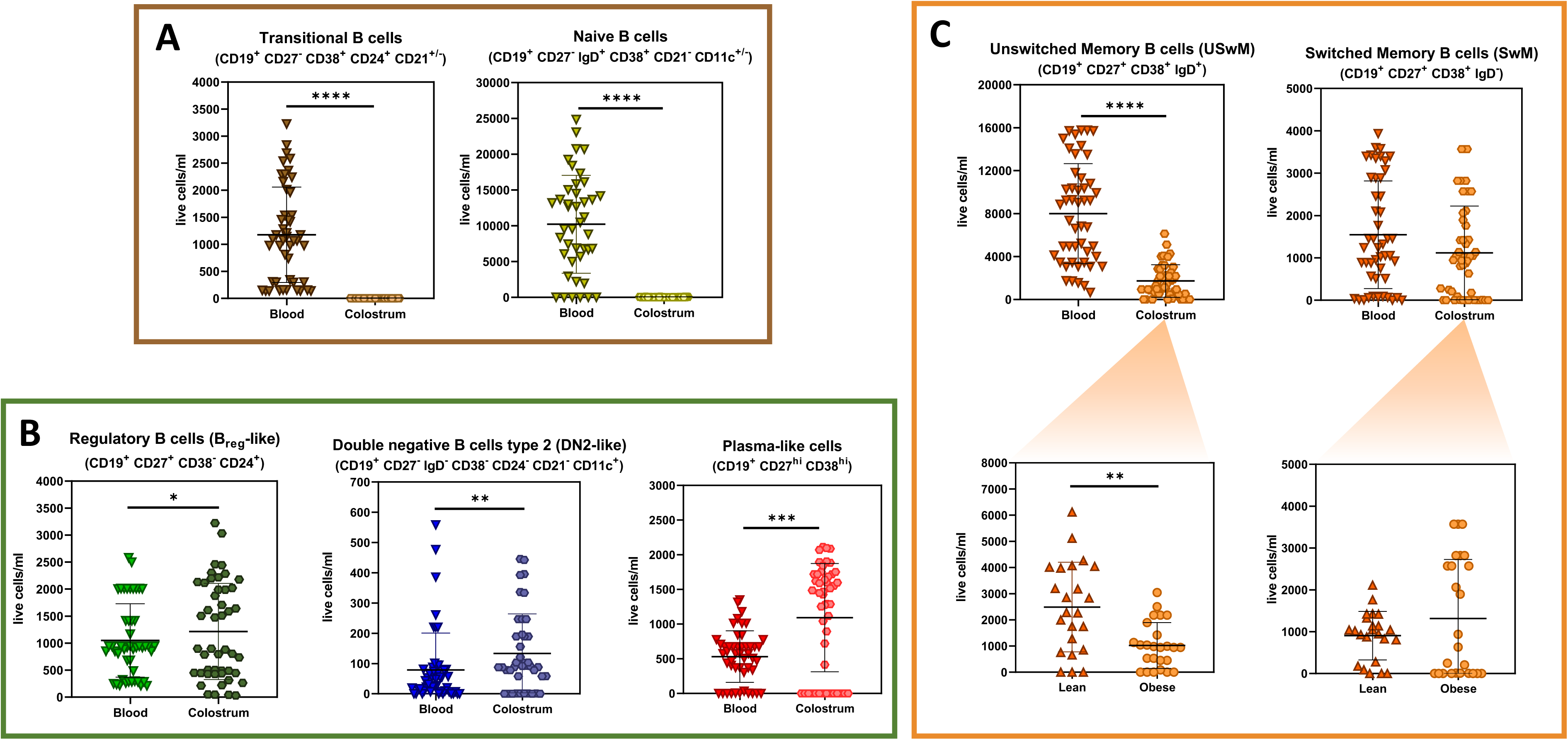
Human colostrum and peripheral blood are differentially enriched in multiple B lymphocyte subtypes. **a)** Concentrations of transitional and naïve B cells in peripheral blood and colostrum. **b)** Concentrations of B_reg_-like, DN2-like and plasma-like cells in peripheral blood and colostrum. **c)** Concentrations of USwM and SwM in blood and colostrum (above) and subsampling comparing samples from mothers with BMI<25 (“lean”) and mothers with BMI>30 (“obese”) (below). Statistical analysis was performed using Mann-Whitney U tests, comparing B cell subsets concentrations (live cells/ml or original blood or colostrum) from mothers with obesity (n=25) or with a lean BMI (n=23). *p < 0.05, **p< 0.01, ***p < 0.001 and ****p < 0.0001.

We then asked if proportions of specific B cell subpopulations in colostrum were regulated with maternal obesity. In this context, we measured significantly less USwM B cells without detected changes in the SwM B cells (Fig. 1C). While this work is the first to report changes in breastmilk memory B cells in relation with maternal obesity, others have measured increased breastmilk SwM B cells from HIV-infected mothers^28^. Increased SwM B cells were also found in mouse visceral adipose tissue^29^. IgG isotype switching follows stimulation with cytokines that are increased in obesity^29,30^. We propose that the increase in SwM B cells depletes USwM, causing the significant decrease in colostrum USwM B cells. In support of this, obesity peripheral blood indeed contained significantly less USwM and significantly more SwM B lymphocytes (data not shown).

### 3.2 Obesity colostrum harbors a dysregulated B lymphocyte repertoire, hinting towards an inflammatory profile

Looking at functional populations, we found a significantly reduced B_reg_-like cell fraction in the colostrum of mothers with obesity (Fig. 2A). The relative abundance of colostrum B_reg_-like cells negatively correlated with maternal pre-pregnancy BMI and current BF%. Inflammation has been linked to decreased B_reg_-like cells functions and growth^31,32^. Obesity is now accepted as a state of chronic inflammation, which supports the physiological interpretation of the results^33,34^. B_reg_ cells limit ongoing immune reactions, restore immune homeostasis, and promote tolerance to commensals of the gut microbiota, suggesting consequences of this reduction for the infant’s pioneering microbiota^35–38^. We then investigated DN2-like B cells, recently described in blood in multiple inflammatory scenarios including autoimmune disorders, acute infections and obesity^22,15,39^. Consistent with the inflammatory state suggested by a reduced B_reg_-like B cell fraction, we found significantly increased proportions of DN2-like cells, and these changes positively correlated with pre-pregnancy BMI and current BF% (Fig. 2B). While the exact origins and roles of DN2-like cells remain unclear to date, their occurrence follow a proinflammatory stimulus^40–43^. Future experiments could include measuring the transcription factor T-bet and IFNγ production from these cells to confirm cell identity^44–46^. We then investigated plasma-like cells that could be producing antibodies. These were increased in obesity colostrum, and as observed with DN2- like B cells, colostrum plasma-like cell proportions positively correlated with pre-pregnancy BMI and with current BF% (Fig. 2C). Plasma cells can differentiate following IFNγ^47^ and leptin signaling^48^. These soluble factors are increased in obesity^49^, which provides a physiological explanation for our findings.

**Fig. 2:**
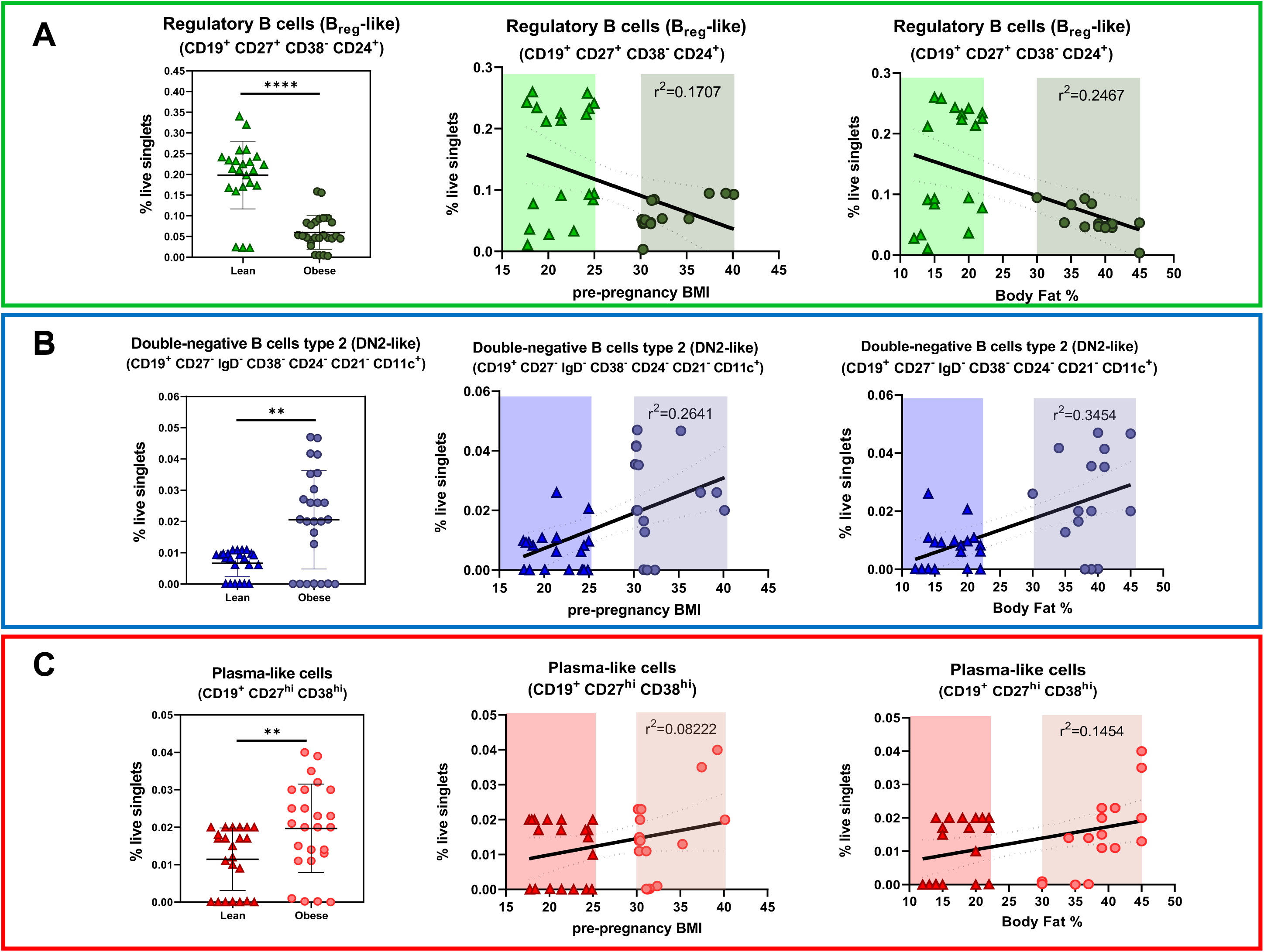
Maternal BMI and BF% correlates with the frequency of Breg-like, double-negative (DN2) and plasma-like cells in colostrum. Comparisons of **a)** B_reg_-like, **b)** DN2-like and **c)** plasma-like cell % in colostrum from lean and obese cohorts (left of the 3 panels), and correlations with between pre-pregnancy BMI values (middle) and BF% (right). Doted lines represent SE. Mann-Whitney U tests were used to compare B cell subsets in colostrum from both cohorts. Pearson correlations were used to investigate the relationships between B cell subsets % and BMI or BF%. **p < 0.01 and ****p < 0.0001.

Correlation values (r^2^) for the 3 cell types were consistently higher for BF% compared to BMI. Despite the historical use of BMI as an indicator of obesity, the lack of precision in the composition of the measured weight is confounding. Our results suggest a clearer association between increased BF% and the regulation of specific B cell subtypes in human colostrum.

### 3.3 Obesity modulates colostrum plasma like-cells and their antibody secreting function

Having identified a significant increase in colostrum plasma-like B cells, we wondered if this population exhibited changes in their antibody production, isotypes, and antigen specificity. As B cells gradually lose CD19 surface expression during differentiation towards antibody-secreting cells, we measured CD19 MFI within the plasma-like cell subpopulations in both groups to compare their relative degree of maturity^50^. We identified 3 discrete subpopulations based on CD19 expression level, with a significant increase in the CD19^low^ plasma-like B cell fraction in the obese cohort (Supplementary Fig. 4)^51^. This suggests obese colostrum is enriched in plasma-like cells maturing towards antibody-secreting cells, possibly driven by proinflammatory signals linked to obesity as described earlier^47,48^.

We compared isotypes of the plasma-like cells and evidenced a significantly increased fraction of IgG^+^ plasma-like cells in obese colostrum, while the IgA^+^ fraction remained unchanged (Fig. 3A). Interestingly, intra-individual correlations of IgA^+^- and IgG^+^-plasma-like cells exhibited a trend whereby obese colostrum contained a switched relation of both isotypes (Fig. 3B), suggesting a compensatory mechanism although more work is required to mechanistically explain this. We then investigated if there were more antibody-secreting cells in obese colostrum. Using an ELISPOT assay, we found a significant increase in IgG-secreting cells in obese colostrum which positively correlated with maternal BMI and BF% (Fig. 3C). While IgA-secreting cell concentrations remained unchanged, we observed a negative correlation between their concentration, and maternal BMI and BF%. These results confirm colostrum contains B cell subsets that actively produce antibodies *in situ*, adding to the current understanding of breastmilk IgG originating from FcN-mediated transcellular translocation^52^. We measured significantly more sIgG and less sIgA concentrations in obese colostrum (Fig. 3D). These changes may affect the establishment of the intestinal microbiota^53^. Dysregulations in maternal antibodies received through breastmilk also impact the growth and maturation of the neonatal intestine^54^. These results confirm previous reports of increased local concentrations of IgG in obesity^55^. We further wondered if the increased IgG may be autoimmune, as accumulating evidence links obesity to autoimmune disorders^56^. We measured the concentration of colostrum IgG specifically recognizing N-acetylglucosamine (GlcNAc), a common bacterial and fungal antigen that bears similarities with circulating hyaluronic acid in obesity^57,58^. We found a very significant increase of anti-GlcNAc IgG in obese colostrum (Fig. 3D). The GlcNAc used as target antigen in the assay was obtained from Group A *Streptococcus pyogenes* (GAS), however the incidence GAS infection is low among pregnant mothers from low income countries like Mexico^59^, and no participant reported GAS infection during pregnancy. These results then suggest autoreactive anti-GlcNAc IgG in obese colostrum, and it will be important to investigate how this affects neonatal gut health.

**Fig. 3:**
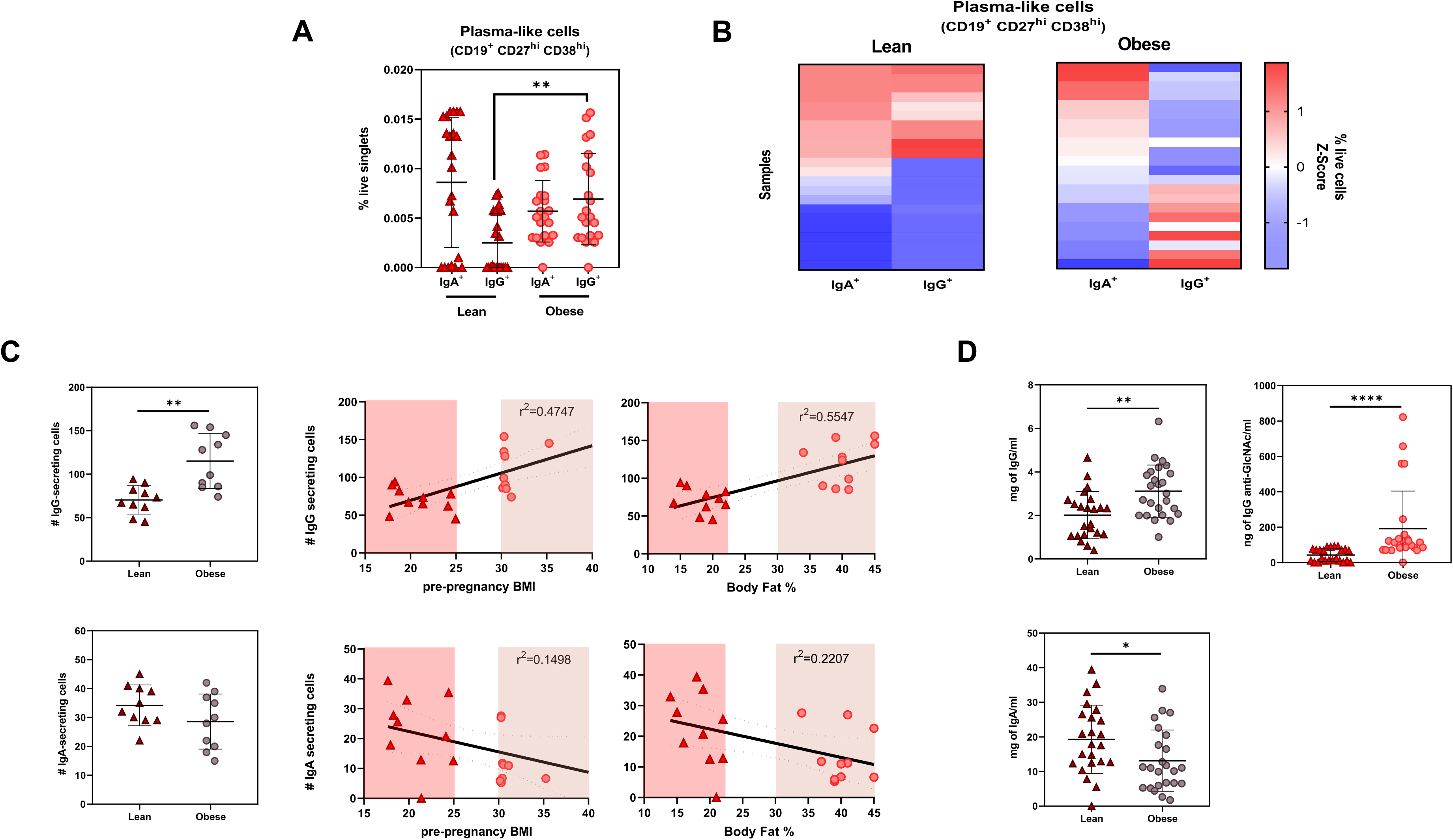
IgG production is increased in colostrum from mothers with obesity. **a)** Relative contribution of IgA^+^ and IgG^+^ plasma-like cells between cohorts (mean ± SD). Comparisons performed using Mann-Whitney U test **p<0.01. **b)** Intra- individual correlation analysis of IgA^+^ and IgG^+^ colostrum plasma-like cells in both groups. Spearman tests compared Z-Scores of IgA^+^ and IgG^+^ plasma-like cells proportions. **c)** Comparison of IgG and IgA-secreting colostrum cells in both cohorts (left), and Pearson correlations with maternal BMI (middle) and BF% (right). **d)** Concentration of total IgG (mg/ml) and GlcNAc-specific IgG (ng/ml) (above) and total IgA (mg/ml) in both groups (individual data, with mean ± SD). Comparisons performed through Mann-Whitney U test *p < 0.05, **p<0.01, ****p < 0.0001.

### 3.4 Obese colostrum IgG may originate from proinflammatory DN2 B lymphocytes

We then wondered what cells could produce IgG in obese colostrum. We noticed DN2-like cells were almost entirely IgG^+^ (Fig. 4A), and these cells have been reported to secrete IgG^40^. Since obese colostrum contained significantly more IgG^+^ DN2-like and IgG^+^ plasma-like cells, together with more sIgG (Fig. 3C) and IgG- secreting cells (Fig. 3A), we correlated IgG concentrations with these possible local producers. There were clear correlations between IgG colostrum concentrations and both IgG^+^ cell types in obesity but not in the lean cohort (Fig. 4B, gray and red data, respectively). Looking at IgG-producing cells, the only significant correlation was with the proportion of DN2-like cells, in obese colostrum only (Fig. 4C, gray data), suggesting these cells participate in the production of IgG in this context. As DN2 cells are mostly found in a proinflammatory setting^15,40,60^, it will be relevant to investigate the IgG subtypes produced and their consequence *in vivo*.

**Fig. 4:**
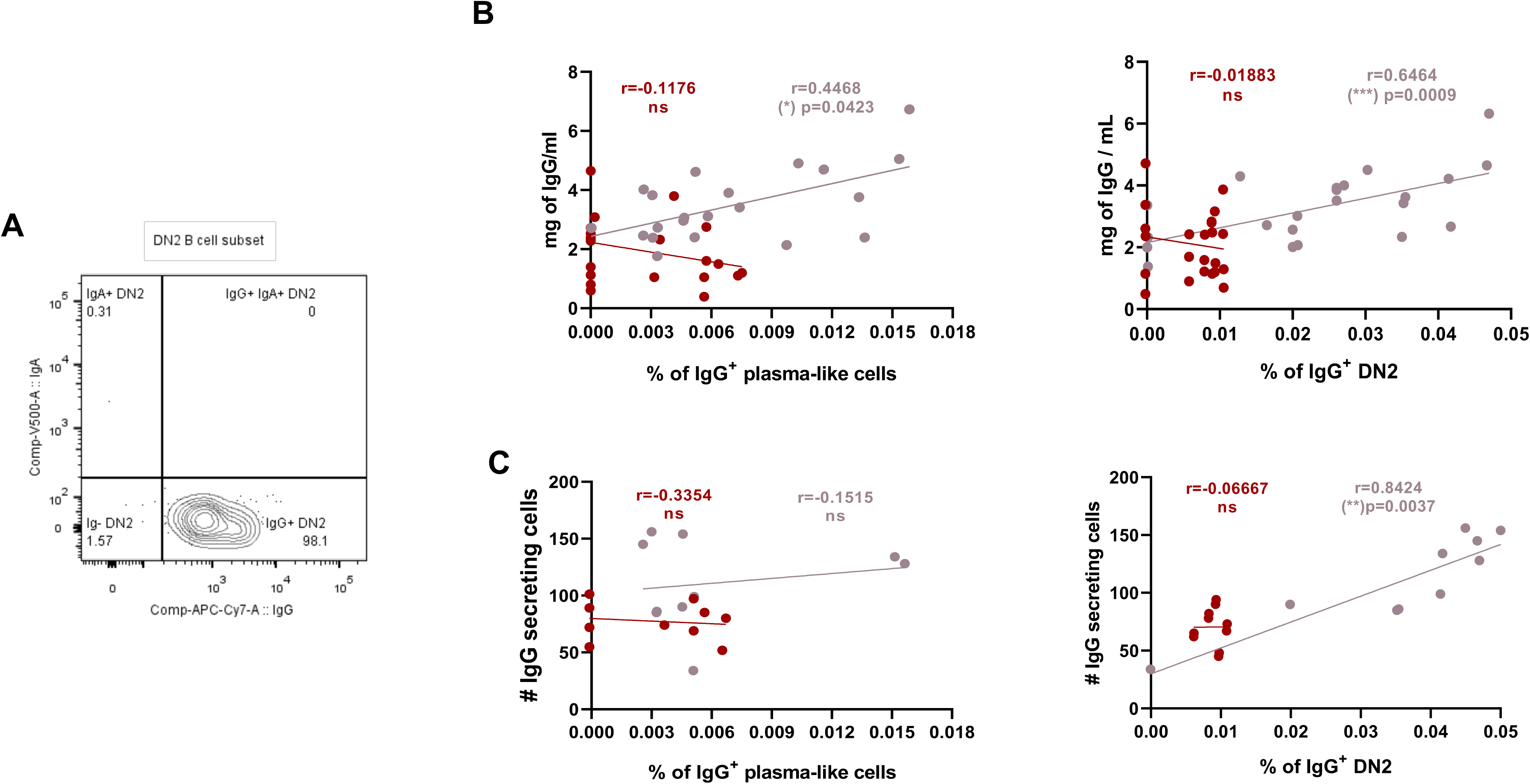
IgG^+^-DN2 cells and IgG^+^-plasma like cells contribute to local IgG production in obesity. **a)** Representative colostrum DN2-like B cells plot showing IgA or IgG expression. **b)** Correlations between IgG^+^ plasma-like cells and DN2- like cell proportions with IgG concentrations in lean (red) and obese (gray) cohorts. **c)** Correlations between IgG^+^ plasma-like cells and DN2-like cell proportions with concentrations of IgG-secreting cells in lean (red) and obese (gray) cohorts. Trends were compared by Spearman rank.

### 3.5 Obese colostrum activates human macrophages *in vitro*

We finally investigated the effect of obese colostrum on human macrophages. Macrophages reside in the neonatal intestine and regulate the local inflammatory response during the first days of life^61^. Co-cultures evidenced that colostrum from mothers with obesity prompted TNF-α production while colostrum from lean mothers did not (Fig. 5A). Until shortly after birth, the neonatal intestine contains macrophages replenished by blood monocytes due to commensal stimulation. In health, these intestinal macrophages show low pro-inflammatory responses, including minimal IL-6 and TNF-α expression^62–66^. Elevated TNF-α levels in the neonatal intestine increases NEC pathogenesis^67^. On the other hand, while IL-6 was significantly increased by obese colostrum stimulation, this cytokine was already present in basal conditions (Fig. 5B). Overall, there was a direct induction of inflammation of macrophages by obese colostrum. Further research should investigate activation mechanisms and long-term consequences for neonatal health.

**Fig. 5:**
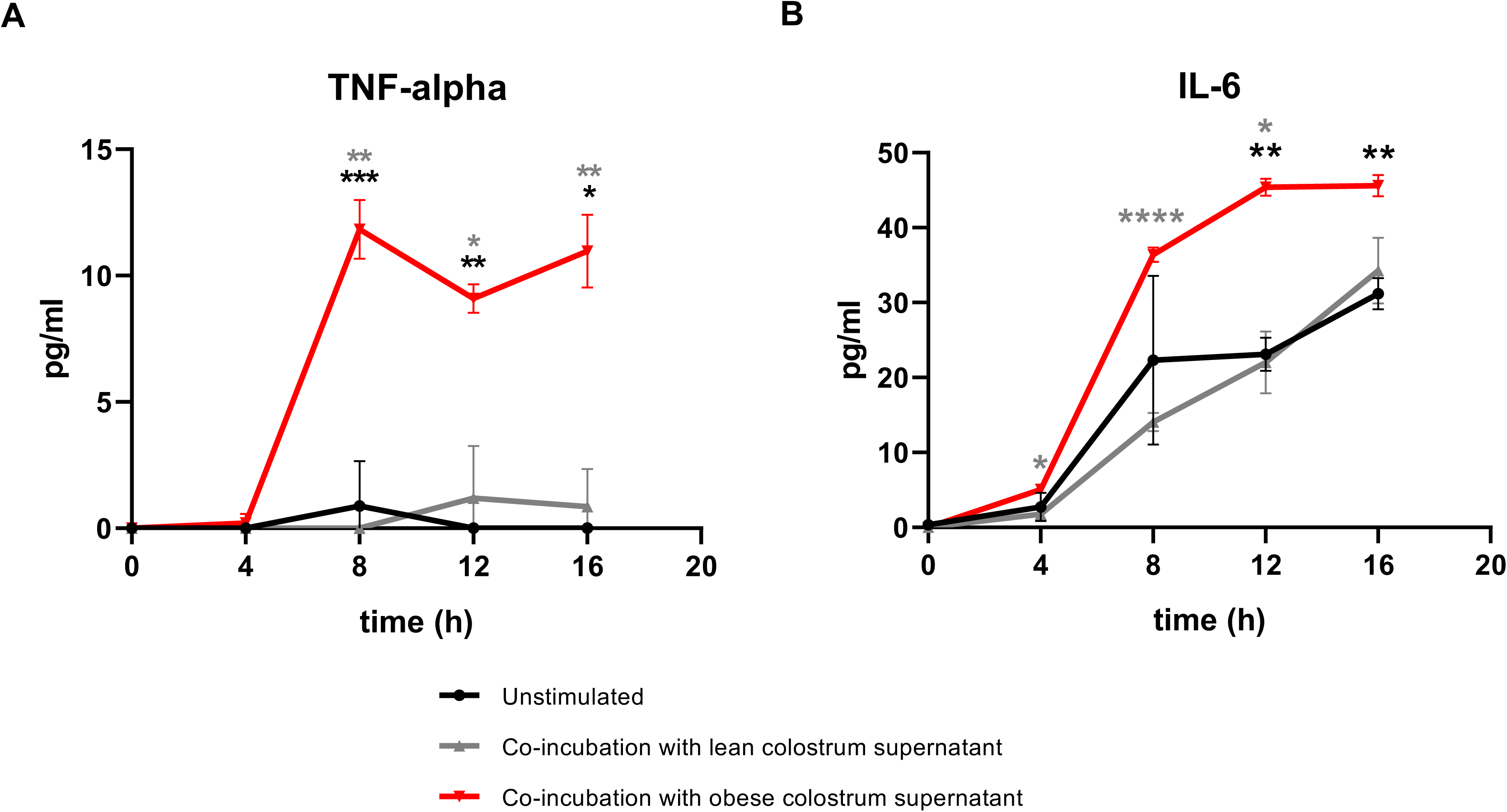
Obese colostrum prompts cytokines production by human macrophages. Supernatant concentrations of **a)** TNF-α and **b)** IL-6 produced over a 16h period by human macrophages (black), human macrophages incubated for 24 h with filtered colostrum supernatant from the lean cohort (gray) or obese cohort (red). Each timepoint represents the mean of three biological replicates ±SD. Comparisons with two-way ANOVA and Turkeýs post-hoc test. *p < 0.05, **p<0.01, ***p<0.001 and ****p < 0.0001.

## 4. Conclusions

This is the first report of obesity-mediated regulation of B lymphocytes and antibodies in human colostrum. We measured notable changes in phenotypically and functionally distinct B lymphocyte subpopulations, which in turn adversely affect the composition of antibodies (summarized in Fig. 6). We advocate for additional research to explore the underlying mechanisms in maternal gut and breast tissue affected by obesity, as well as to understand the ramifications for neonatal intestinal maturation, including the establishment of gut microbiota and maturation of the intestine in suckling infants.

**Fig. 6:**
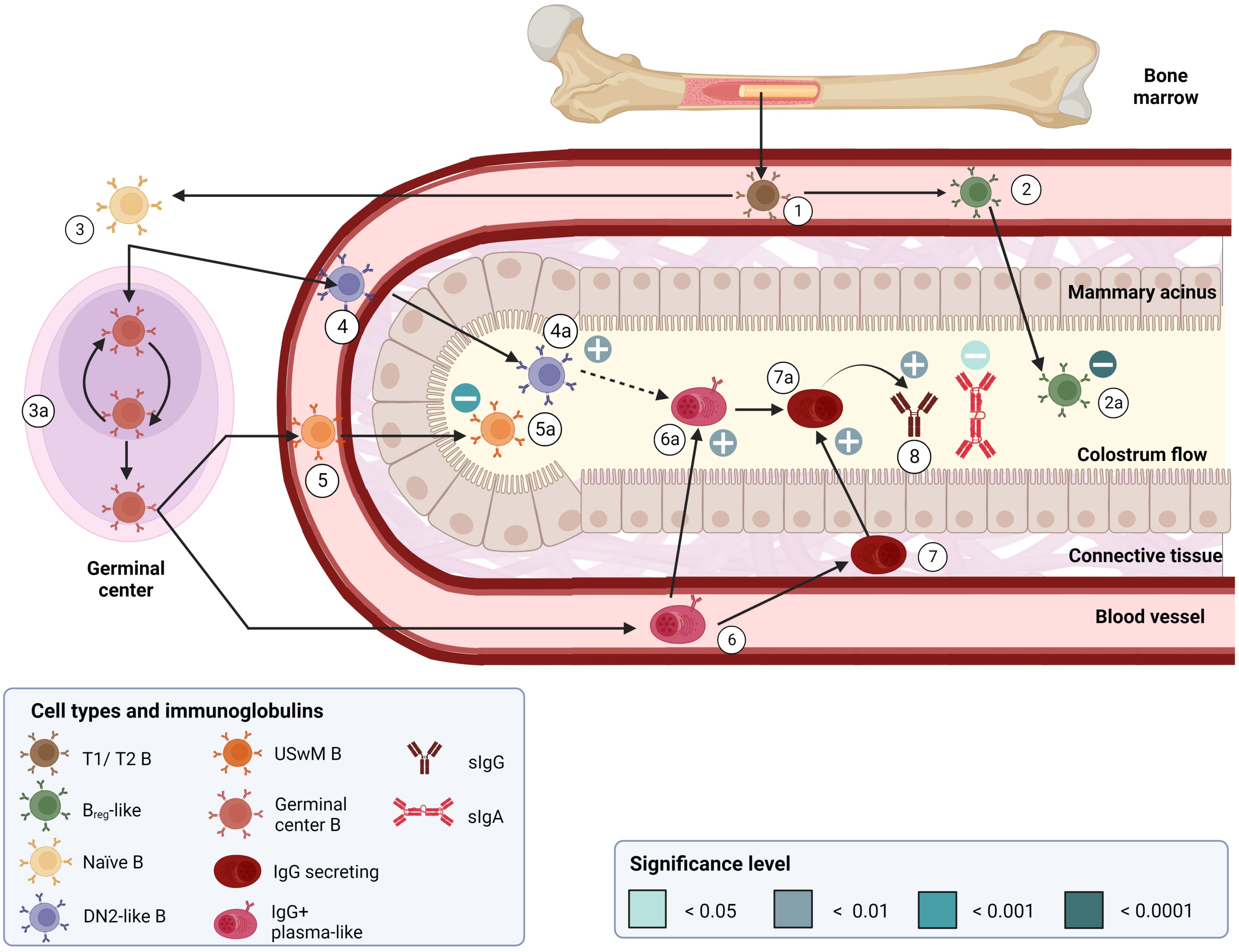
Overview of obesity-related alterations in B lymphocyte subtypes in maternal peripheral blood and colostrum. Transitional B cells (1) differentiate into B_reg_ cells (2) and migrate from the blood into the lactating duct (2a). Transitional B cells (1) also migrate to secondary tissues. There, transitional B cells differentiate into naïve B cells (3) and enter germinal centers (3a) or are activated through the extrafollicular pathway to differentiate into DN2-like B cells (4). These (4) could migrate to multiple tissues, including the lactating mammary gland and colostrum (4a), where they are increased in obesity. Here we provide evidence that DN2-like B cells could differentiate into IgG^+^ plasma-like cells (6a) and IgG- secreting cells (7a) to increase IgG produced in obesity (8). In the germinal center, naïve B cells (3), after germinal center reaction (3a) and can differentiate into two subpopulations: (5) memory B cells or (6) plasma-like cells. Memory B cells migrate through blood to mammary acini and colostrum (5a). We have shown that in obesity, colostrum USwM cells are decreased. Finally, colostrum IgG-secreting cells and sIgG increase in obesity (7a) while colostrum IgA-secreting cells and sIgA are reduced (8). Created with BioRender.com

## AUTHOR CONTRIBUTIONS

All authors approved the final version of the manuscript. ESS designed and performed experiments, analyzed data and wrote, edited, and reviewed the manuscript. DBR performed experiments, analyzed data and reviewed the manuscript. MRAG, VJLD and CNLV, designed experiments, enrolled participants, collected samples, edited, and reviewed the manuscript. MEGB designed the study, analyzed data, wrote, edited, and reviewed the manuscript.

## DATA AVAILABILITY

All .fcs3 files obtained as part of this work will be made freely available on FlowRepository upon paper acceptance (in process)

## ACKNOWLEDGEMENTS

We acknowledge institutional support in all administrative processes involved with a clinical study. We are deeply grateful to participating families.

---

**Supplementary Fig. 1:**
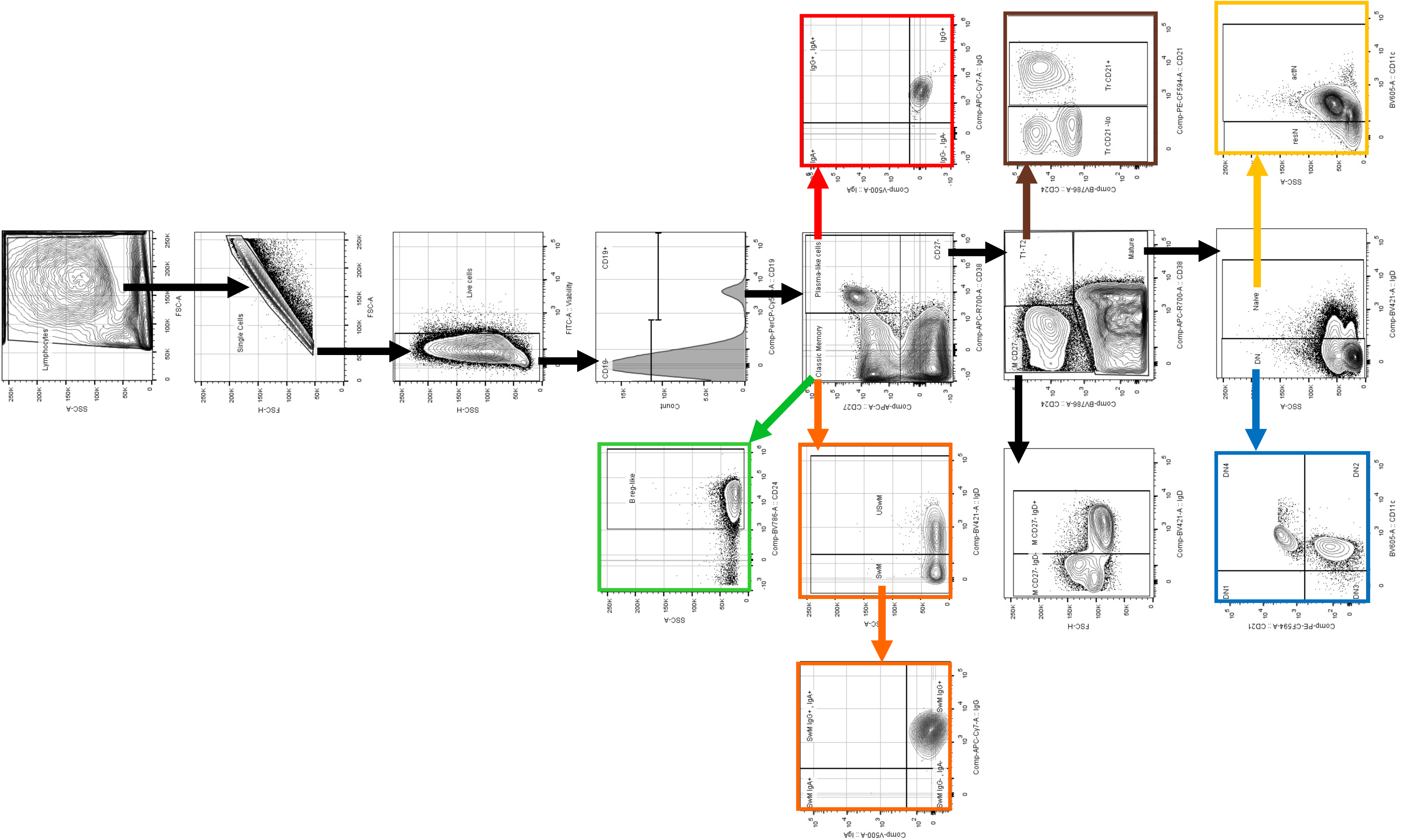
Gating strategy used for manual gating of B cells subsets in colostrum. Transitional B cells in brown, naïve B cells in yellow, regulatory B cells in green, memory B cells in orange, plasma-like cells in red, and double-negative B cell subsets (DN1, DN2, DN3 and DN4) in blue. Dot plots are represented in Contour plots, at least at 5% of level and included outliers in FlowJo X® version Software, BD Biosciences.

**Supplementary Fig. 2:**
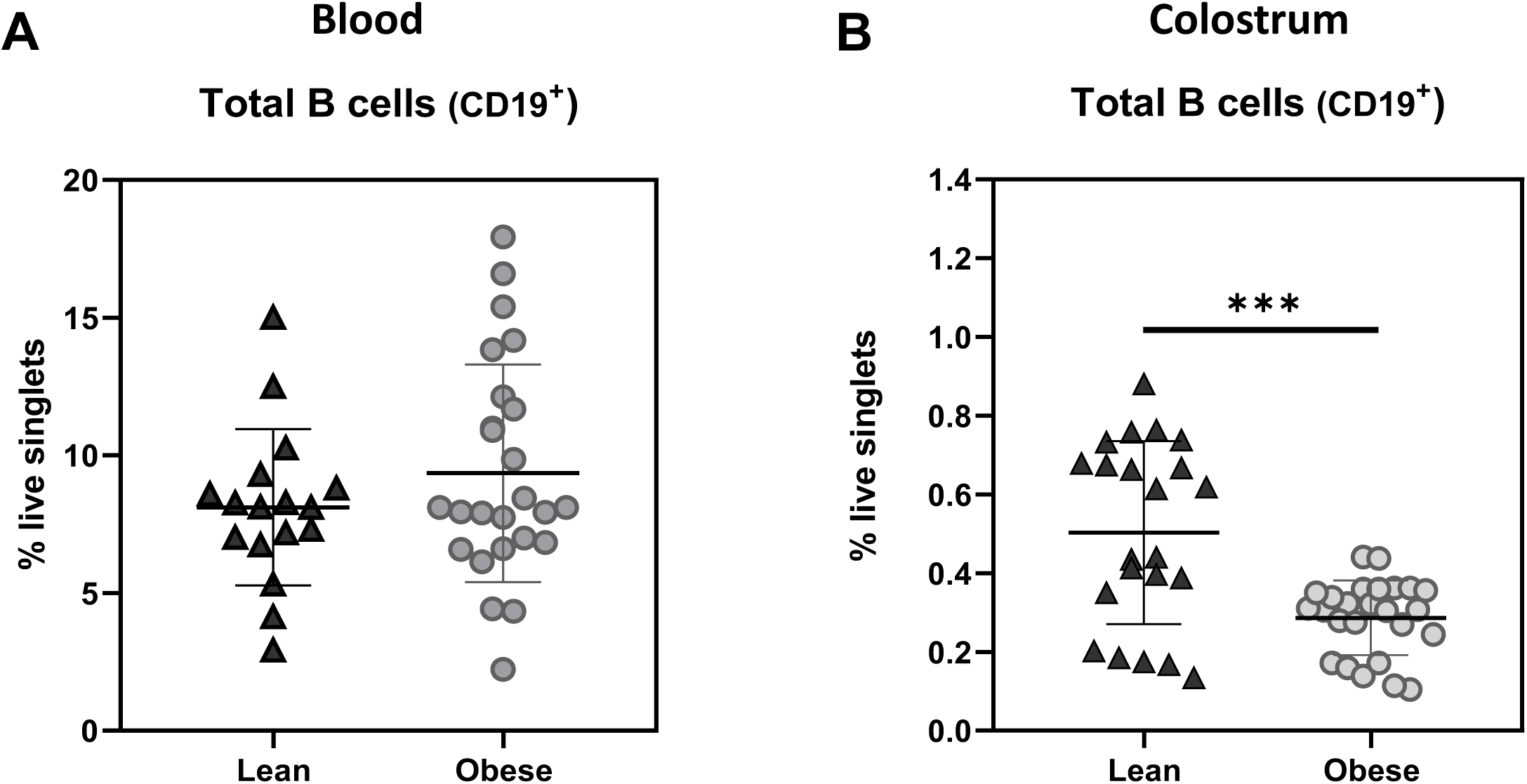
CD19^+^ B cells are decreased in the colostrum from mothers with obesity. **a)** Total CD19^+^ B cells in blood and in colostrum from both cohorts. Groups compared using Mann-Whitney U test. ***p < 0.001.

**Supplementary Fig. 3:**
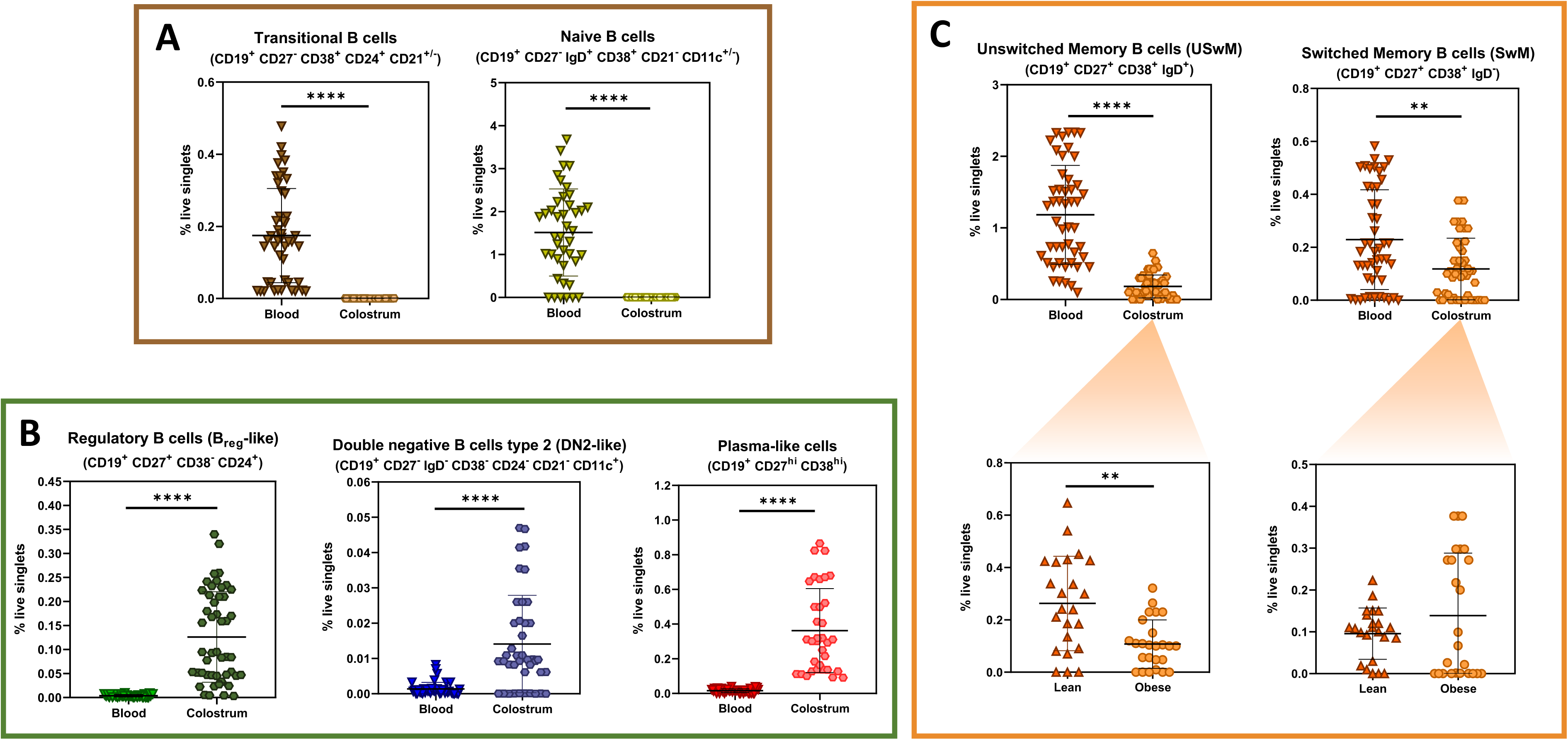
Human colostrum contains percentages of subpopulations of differentiated B cells enriching in comparison with blood. **a)** Scatter plots comparing transitional and naïve B cells in blood and colostrum. **b)** Scatter plots comparing regulatory B cells (Breg-like), double negative 2 (DN2-like) and plasma-like cells in blood and in colostrum. **c)** Scatter plots comparing unswitched memory (USwM) and switched memory B cells (SwM) in blood and colostrum (above). Scatter plots comparing USwM and SwM in colostrum from lean and mothers with obesity (below). Statistical analysis was performed using the Mann-Whitney U test, comparing B cell subsets percentages in blood from mothers with obesity (n=25) against lean subjects as control (n=23). *p < 0.05, **p< 0.01, ***p < 0.001 and ****p < 0.0001.

**Supplementary Fig. 4.**
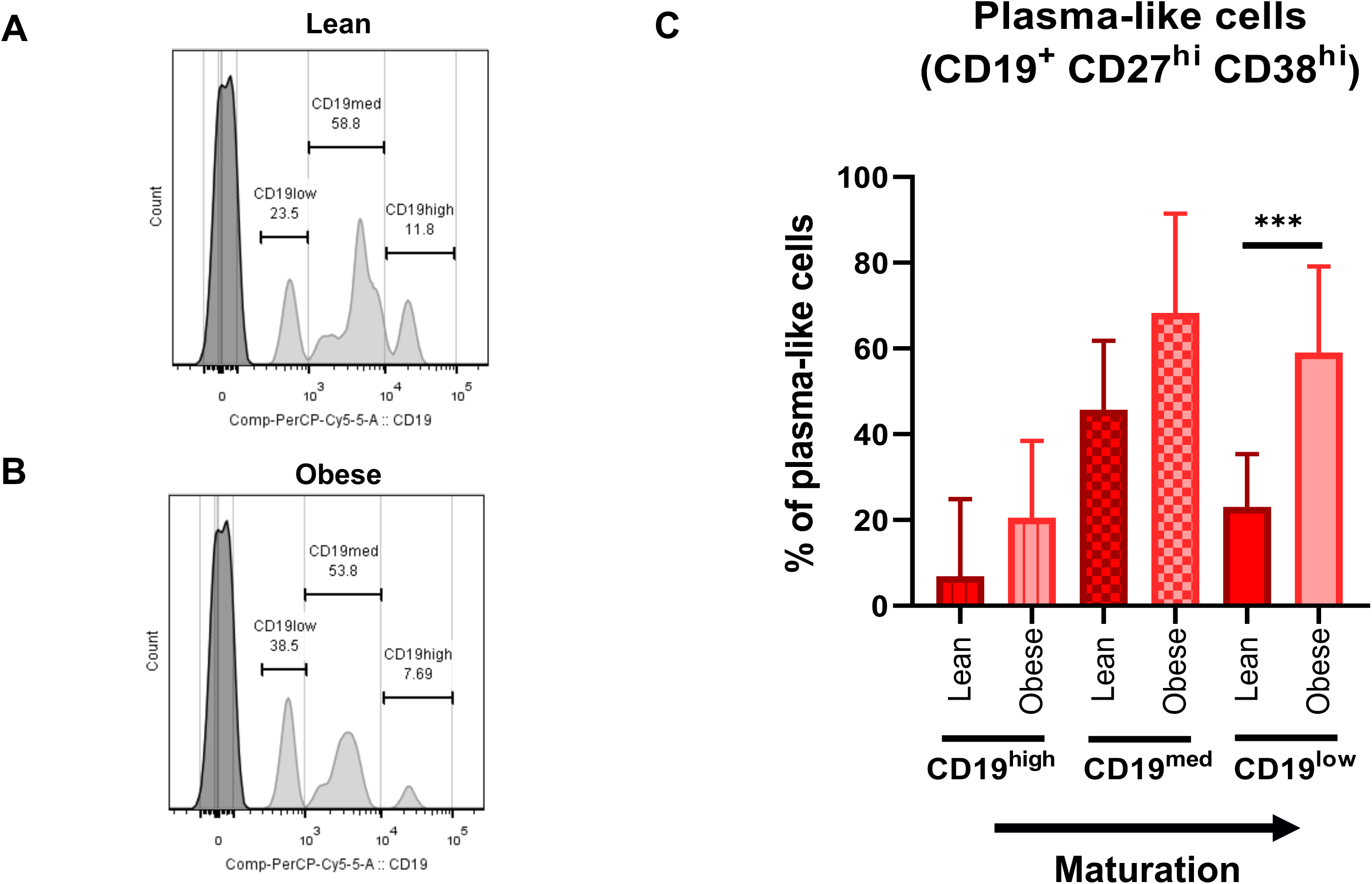
Obesity induces differentiation of plasma-like cells on CD19 low expression group. **a,b)** Examples of histograms from plasma-like cells analyzed in function of CD19 expression, showing negative CD19 population (left) and positive CD19 expression groups (right), from colostrum of lean and mothers. **c)** Column bar graph comparing plasma-like cells in function of percentage of (% of CD19^+^) low (no pattern), median (square pattern), and high (line pattern) CD19 expression groups. Bars indicate the mean ± SD. Statistical analysis was performed using the Kruskal Wallis test for multiple comparisons (Lean n=23 and obesity n=25) assay. ***p<0.001.

